# Identification of IgG antibody response to SARS-CoV-2 spike protein and its receptor binding domain does not predict rapid recovery from COVID-19

**DOI:** 10.1101/2020.05.01.20087684

**Authors:** Kathleen M. McAndrews, Dara P. Dowlatshahi, Janine Hensel, Luis L. Ostrosky-Zeichner, Ramesh Papanna, Valerie S. LeBleu, Raghu Kalluri

## Abstract

Diagnostic testing and evaluation of patient immunity against the novel severe acute respiratory syndrome (SARS) corona virus that emerged last year (SARS-CoV-2) are essential for health and economic crisis recovery of the world. It is suggested that potential acquired immunity against SARS-CoV-2 from prior exposure may be determined by detecting the presence of circulating IgG antibodies against viral antigens, such as the spike glycoprotein and its receptor binding domain (RBD). Testing our asymptomatic population for evidence of COVID-19 immunity would also offer valuable epidemiologic data to aid health care policies and health care management. Currently, there are over 100 antibody tests that are being used around the world without approval from the FDA or similar regulatory bodies, and they are mostly for rapid and qualitative assessment, with different degrees of error rates. ELISA-based testing for sensitive and rigorous quantitative assessment of SARS-CoV-2 antibodies can potentially offer mechanistic insights into the COVID-19 disease and aid communities uniquely challenged by limited financial resources and access to commercial testing products. Employing recombinant SARS-CoV-2 RBD and spike protein generated in the laboratory, we devised a quantitative ELISA for the detection of circulating serum antibodies. Serum from twenty SARS-CoV-2 RT-PCR confirmed COVID-19 hospitalized patients were used to detect circulating IgG titers against SARS-CoV-2 spike protein and RBD. Quantitative detection of IgG antibodies to the spike glycoprotein or the RBD in patient samples was not always associated with faster recovery, compared to patients with borderline antibody response to the RBD. One patient who did not develop antibodies to the RBD completely recovered from COVID-19. In surveying 99 healthy donor samples (procured between 2017-February 2020), we detected RBD antibodies in one donor from February 2020 collection with three others exhibiting antibodies to the spike protein but not the RBD. Collectively, our study suggests that more rigorous and quantitative analysis, employing large scale samples sets, is required to determine whether antibodies to SARS-CoV-2 spike protein or RBD is associated with protection from COVID-19 disease. It is also conceivable that humoral response to SARS-CoV-2 spike protein or RBD works in association with adaptive T cell response to determine clinical sequela and severity of COVID-19 disease.

## Introduction

The COVID-19 pandemic, a respiratory illness caused by infection with the novel corona virus SARS-CoV-2, is a rampant health crisis that also severely impacted the financial security and access to care of many, in particular our most vulnerable communities^1,2^. As social distancing measures appear to reduce the spread of the virus, the development of testing modalities for diagnosis and acquired immunity for SARS-CoV-2 are essential to inform health care management and public health policies. Lessons from previous corona virus infection studies, including the moderate upper respiratory tract disease or ‘common cold’ caused by the human corona viruses, CoV-229E, CoV-NL63, CoV-OC43, or CoV-HKU1, and the more severe respiratory disease caused by SARS-CoV or MERS-CoV, support an idea that prior exposure could result in immunity to the virus, which protects against re-infection and may limit symptoms if re-infection occurs^3,4^. Reports on immunity (detection of antibodies) against corona virus (mainly SARS-CoV) acquired from exposure indicate circulating antibodies are observed from 2 to 17 years following recovery^5,6^.

Data from China, the epicenter of COVID-19, indicated up to 27% of SARS-CoV-2 patients 85 years or older die from the disease, in contrast with approximately 1% of SARS-CoV-2 patients 54 years or younger^7^. Recent findings also highlight age as a significant factor associated with mortality and hospitalization in Europe and the USA due to COVID-19^8,9^, with 21% mortality observed in 56 to 78 years old New Yorkers^9^. This may be reflecting aging-associated changes in the adaptive immune system^10^, and possibly other co-morbidities, including hypertension, obesity, and diabetes^9^ which could also play a role in altering immune response to infection. The activation of the immune system in response to SARS-CoV-2 infection and clinical presentation is likely complex and further studies are required to measure immune responses towards infection, recovery, and development of immunity. Emerging findings mostly rely on confirmed or suspect symptomatic cases receiving care at health care facilities, and it remains unclear today what percentage of the population has been exposed to SARS-CoV-2 and remained asymptomatic, or mildly symptomatic, since they did not require care and thus were not captured in healthcare records. Such unaccounted cases could underestimate the reported percentage of the population that has been exposed to SARS-CoV-2. This limitation also challenges the ability for precise assessment of the percentage of our population that has developed immunity to SARS-CoV-2. The detection of circulating antibodies against SARS-CoV-2 may inform on immunity to the virus, and ongoing efforts for sensitive and specific assays include the development of lateral flow chromatographic immunoassay (LFIA) or enzyme-linked immunoabsorbent assays (ELISA) designed to detect antibodies against SARS-Cov-2 viral proteins, some of which were granted emergency use authorization by the FDA (https://www.fda.gov). Additional validation studies are critically needed to define the utility of the assays under development, and consideration vis-a-vis cost and accessibility must be taken in consideration to enable testing in communities that require the most assistance. To this end, we developed and tested a simple procedure to enable the detection of circulating antibodies against SARS-Cov-2 viral proteins.

SARS-CoV-2 is an enveloped (+) ssRNA virus that encodes 14 viral proteins, including structural proteins, a replicase, and proteases. Viral proteins on the surface of the SARS-CoV-2 envelope interact with human cells to promote entry of the virus into the cell. Specifically, the binding of the viral surface spike (S) glycoprotein to the angiotensin converting enzyme (ACE2) on human lung epithelial cells enables internalization via receptor mediated endocytosis, a potentially rate-limiting step for infectivity and viral propagation^11,12^. The receptor binding domain (RBD) of S protein interact with ACE2 with high affinity^13^. The S protein and its RBD emerged as potential antigens against which humoral immunity may develop, and the role of the S protein and its RBD in viral entry suggest antibodies against these proteins may present with neutralizing properties. Monoclonal antibodies cloned from memory B cells of convalescent COVID-19 patients demonstrated S protein binding and in vitro neutralizing activity^14^. Seroconversion yielding circulating IgG antibodies against such antigen may thus inform on acquired SARS-CoV-2 immunity. Notably, studies of circulating antibodies from patients with SARS-CoV and SARS-CoV-2 showed little cross-neutralization^15^, supporting that antibodies against the S protein of SARS-CoV-2 may be specific to the novel virus. In 3 of COVID-19 patients serum, antibodies to both RBD and spike protein of SARS-CoV-2 were detected by ELISA when compared to serum from patients with acute NL63 infection, and convalescent serum following dengue, chikungunya, and hantavirus infection^16^, and in a study of 175 adult Chinese COVID-19 convalescent patients, circulating IgG antibodies did not cross react with SARS-CoV antigen^17^. In the latter, the specificity and sensitivity of the ELISA assays for RBD, S1, S2 subunits of the SARS-CoV-2 spike protein was however not specifically reported^17^. These studies informed on the feasibility of detecting circulating antibodies to SARS-CoV-2 with the potential to inform on acquired immunity, and support diagnosis. The detection of circulating antibodies for example offered added sensitivity to RNA detection (RT-PCR assay) for the diagnosis of COVID-19 in 173 hospitalized Chinese patients^18^. A comparative analysis of LFIA and ELISA for SARS-CoV-2 full length spike protein IgG and IgM antibodies indicated ELISA was superior for detection of antibodies in 40 COVID-19 patients and 50 pre-2019 donors, and SARS-CoV-2 spike protein IgG antibodies were detected in all 31 RT-PCR COVID-19 positive patients 10 days after diagnosis and up to 8 weeks post diagnosis^19^. In 16 European RT-PCR confirmed COVID-19 patients, ELISA captured circulating IgG antibodies against SARS-CoV-2 spike protein when compared to control samples as early as 5 days following onset of symptoms, and plaque-reduction neutralization tests support such antibodies have neutralizing properties^20^. Building upon the recent findings that unveiled the structure of the SARS-CoV-2 spike protein and its binding with human ACE2^21^ and the emerging data on the early detection of potentially neutralizing antibodies binding to SARS-CoV-2 spike protein, we generated and tested SARS-CoV-2 spike protein and RBD as antigen substrates. The substrates were utilized in ELISAs designed to detect IgG antibodies against SARS-CoV-2 spike protein and RBD. We ascertained SARS-CoV-2 IgG antibody levels in 99 healthy donor serum samples collected from 2017 to 2020, as well as serum from 20 RT-PCR validated SARS-CoV-2 positive COVID-19 patients.

## Methods

### Human serum

All human serum samples (n=99) were obtained under IRB exempt and were de-identified. Frozen serum from healthy donors were obtained from the MD Anderson Laboratory Medicine at MD Anderson Cancer Center, and donations spanned from 2017 to 2020. Serum from SARS-CoV-2 patients were obtained from the Memorial Hermann Hospital-Texas Medical Center (n=20) under the approval of Institutional Review board at the University of Texas McGovern Medical School at Houston. Patients characteristics are described in **Table 1**. All samples were heat inactivated at 56°C for 30 minutes and stored at −80°C until assayed as described therein.

**Table 1.**
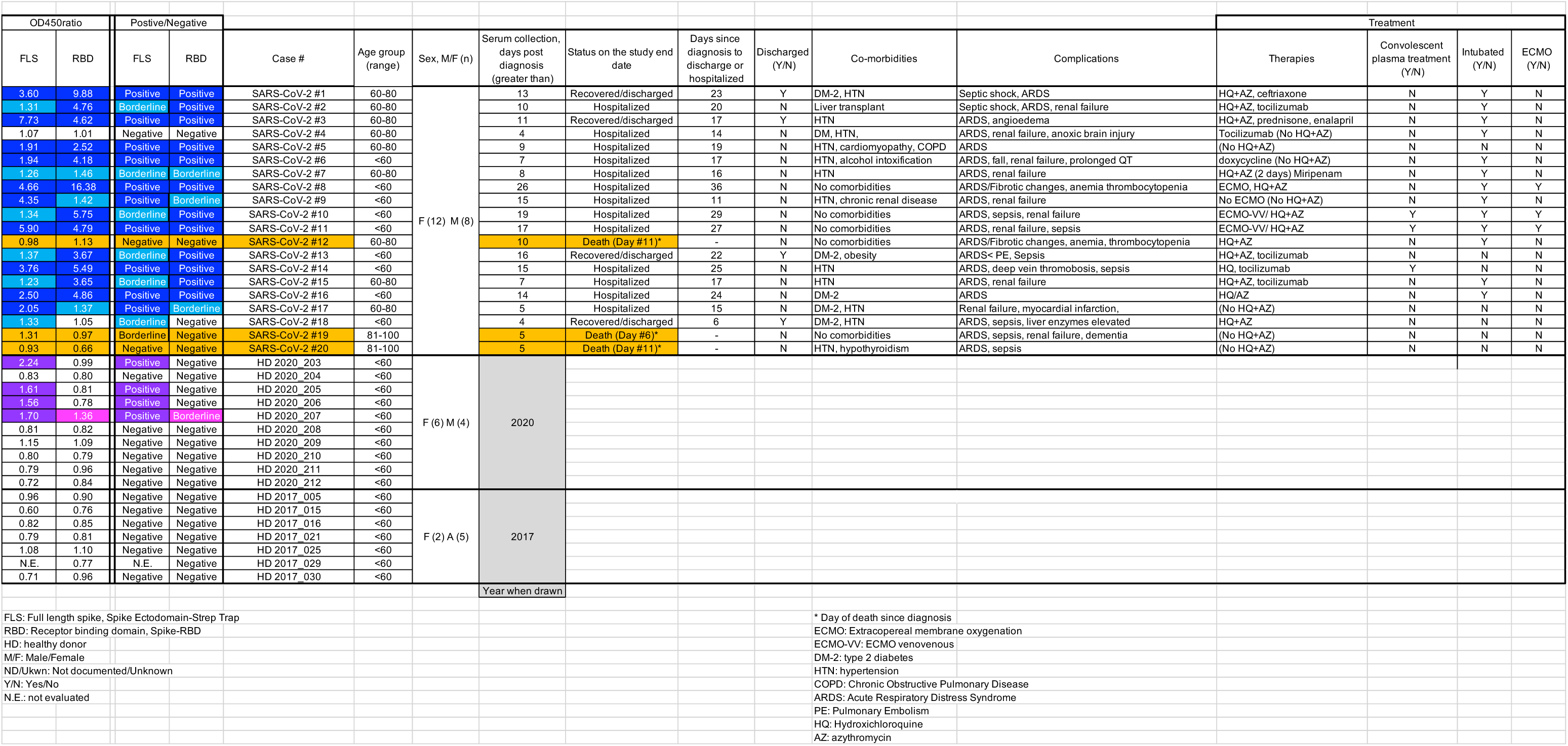
Patient Characteristics.

### Plasmid

The SARS-CoV-2 spike protein ectodomain expression plasmid (**Figures 1b**) were a kind gift from Dr. McLellan at the University of Texas, Austin, TX, and was previously detailed^21^. Transformation of *E. Coli* (TOP10 competent cells, Invitrogen) cultured under ampicillin selection enabled plasmid production, which was purified using NucleoBond Xtra Maxi EF (Macherey-Nagel) according to the manufacturer’s directions. Restriction digestion of the purified plasmid (**Figure S1b**) utilized enzymes purchased from New England Biolabs and restriction digestions were carried out as recommended by the manufacturer. Digestion products were visualized following electrophoresis on a 0.7% agarose gel. The spike-RBD plasmids were obtained from Sino Biological (VG40592-CH) or ATUM (SARS-CoV-2-spike RBD chimera to VSV-G transmembrane domain).

**Figure 1:**
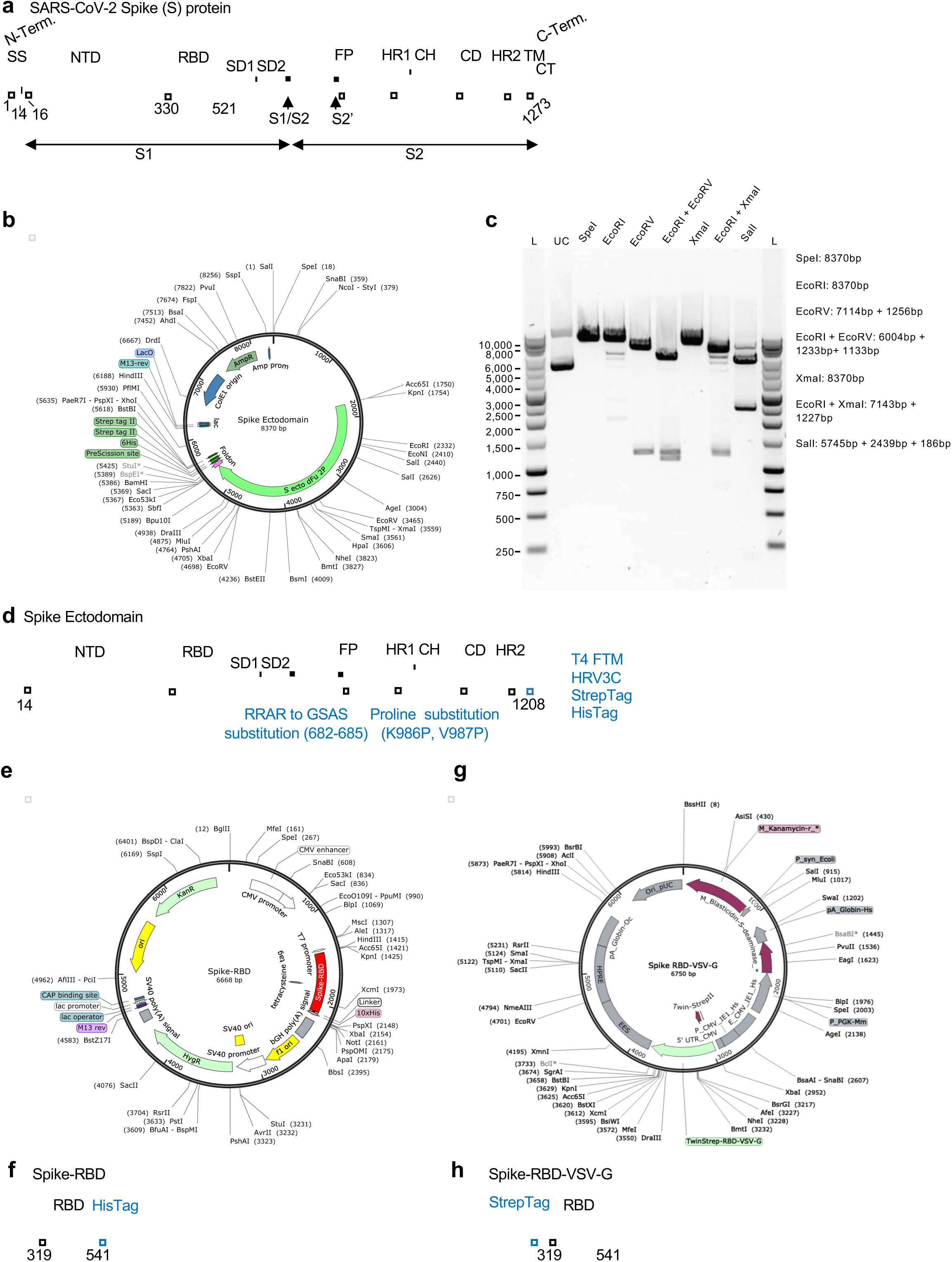
Plasmids used for generation of recombinant SARS-CoV-2 spike protein. (**a**) Schematic of SARS-CoV-2 spike protein. SS, signal sequence; NTD, N-terminal domain; RBD, receptor binding domain; S2’, S2’ protease cleavage site; FP, fusion peptide; HR1, heptad repeat 1; CH, central helix; CD, connector domain; HR2: heptad repeat 2; TM, transmembrane domain; CT, C-terminus. (**b**) Map of SARS-CoV-2 spike protein ectodomain expression plasmid construct (Spike Ectodomain) used to generate Spike Ectodomain protein. (**c**) Agarose gel electrophoresis of restriction digested Spike Ectodomain plasmid. L: ladder, UC: uncut plasmid. Predicted sizes of each digestion denoted. (**d**) Schematic of Spike Ectodomain protein. NTD, N-terminal domain; RBD, receptor binding domain; FP, fusion peptide; HR1, heptad repeat 1; CH, central helix; CD, connector domain; HR2: heptad repeat 2; T4 FTM, T4 fibritin trimerization motif; HRV3C, HRV 3C protease cleavage site. Protein modifications are indicated in blue. (**e**) Map of SARS-CoV-2 spike protein RBD expression plasmid construct (Spike-RBD). (**f**) Schematic of Spike-RBD protein. (**g**) Map of SARS-CoV-2 spike protein RBD expression plasmid construct (Spike-RBD). (**h**) Schematic of Spike-RBD-VSV-G protein. RBD, receptor binding domain; VSV-G, vesicular stomatitis virus G.

### Cell culture and transfection

293T/17 (HEK 293T/17) cells from ATCC (CRL-11268) were cultured in DMEM (Corning) with 10% fetal bovine serum (FBS, Gemini). The cells were transfected with the listed plasmids using lipofectamine 3000 (Invitrogen) according the manufacturer’s protocol, and the cells were maintained in DMEM supplemented with 10% FBS. Conditioned media (CM) was collected for two consecutive 48 hours incubation (CM1 and CM2, respectively, **Figure 2a**).

**Figure 2:**
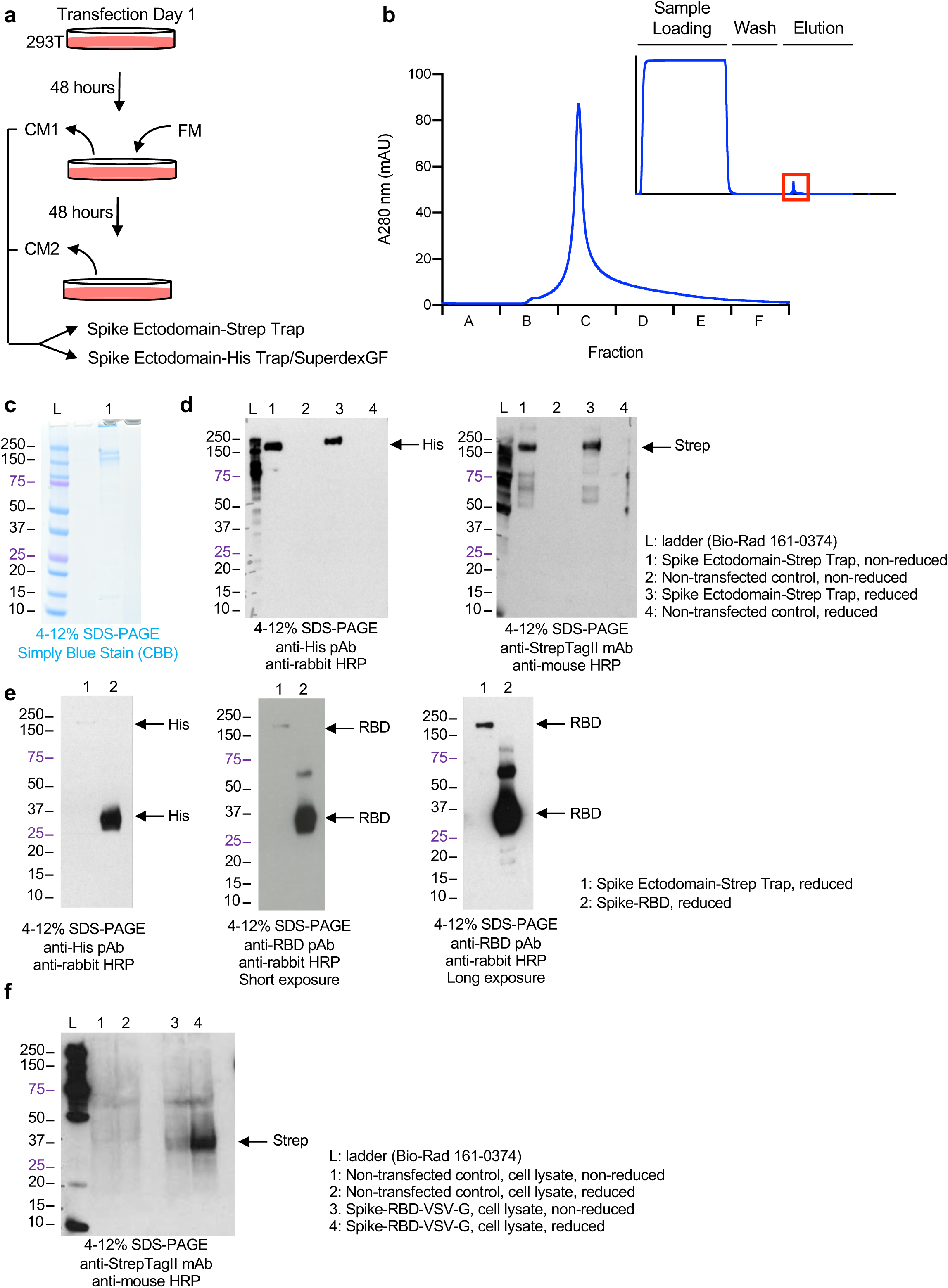
Generation and validation of recombinant SARS-CoV-2 spike protein. (**a**) Schematic of conditioned media collection of Spike Ectodomain transfected 293T cells. Cells are transfected, conditioned media collected 48 hours later (CM1), fresh media (FM) added, and conditioned media collected 48 hours later (CM2). (**b**) Representative chromatogram of Strep-Trap isolated Spike Ectodomain. (**c**) Representative Coomassie stain of Strep-Trap purified Spike Ectodomain. (**d**) Representative Western blots for His-tag (left panel) and Strep-tag (right panel) of Strep-Trap purified Spike Ectodomain. (**e**) Representative Western blot for His-tag (left panel) and spike protein RBD (long exposure, middle panel; short exposure, right panel) of Strep-Trap purified Spike Ectodomain and Spike-RBD. (**f**) Representative Western blot for Strep-tag of Spike-RBD-VSV-G transfected 293T cells.

### Protein purification and quantification

The conditioned media from transfected 293T/17 cells was collected, centrifuged at 800 × g for 5 minutes, filtered with a 0.22 μm filter (ThermoScientific), and stored at −80°C prior to processing. Affinity purification was performed using an Akta Pure fast protein liquid chromatography (FPLC) system (Cytiva, formerly GE Healthcare), and size exclusion chromatography (SEC) was performed using a high-performance liquid chromatography (HPLC) system (Agilent 1200). CM was subjected to two distinct purification methods, yielding Spike Ectodomain-Strep Trap and Spike Ectodomain-His Trap/SuperdexGF products, respectively. The Spike Ectodomain-Strep Trap product purification was performed using StrepTactin Sepharose High Performance resin, where samples were supplemented to a final concentration of 1x EDTA-free protease inhibitor cocktail (Roche) before loading onto a StrepTrap HP 1mL column (Cytiva, formerly GE Healthcare, equilibrated in PBS 300 mM NaCl, pH 7.4), and eluted over six column volumes of elution buffer (PBS 300 mM NaCl, 2.5 mM Biotin). Eluted fractions were pooled, buffer exchanged into PBS with 30,000 Da molecular weight cutoff (MWCO) ultrafiltration membrane spin column (Amicon) per the manufacturer’s instructions, and stored at −80°C.

The Spike Ectodomain-His Trap/SuperdexGF product purification was performed using nickel-nitrilotriacetic acid (NiNTA) resin, where samples were supplemented to a final concentration of 1 mM Imidazole (Sigma) and 1x EDTA-free protease inhibitor cocktail (Roche) before loading onto a Qiagen NiNTA Superflow 5 ml column (Cytiva, formerly GE Healthcare) equilibrated in wash buffer (PBS 300 mM NaCl, 10 mM Imidazole, pH 8.0), and eluted with a linear gradient from 10 to 500 mM Imidazole. Eluted fractions were pooled, concentrated with 30,000 Da (MWCO) ultrafiltration membrane spin column (Amicon) per the manufacturer’s instructions, and purified by SEC on a Superdex 200 Increase 10/300 GL column (Cytiva, formerly

GE Biosciences) equilibrated with PBS, pH 7.4). Fractions from SEC were pooled, concentrated with 30,000 Da MWCO ultrafiltration membrane spin column (Amicon) and stored in PBS at −80°C. The list of reagents for the protein purifications are listed in **Table S1**. The purified spike ectodomain proteins were quantified using NanoDrop OneC (280 nm absorbance, Thermo Scientific), and BCA assay (Thermo Scientific) relative to bovine serum albumin (BSA) standard according to manufacturer’s instructions.

### Western blot

Spike Ectodomain-Strep Trap, Spike Ectodomain-His Trap/SuperdexGF, and Spike-RBD-VSV-G were electrophoresed on 4-12% SDS-PAGE gel (NOVEX) stained with SimplyBlue SafeStain (Invitrogen). For immunolabeling, the proteins were transferred onto 0.45 um PVDF membrane (Bio-Rad) using a Trans-blot Turbo Transfer System (Bio-Rad). Cell lysates were collected in RIPA buffer with protease inhibitor (Roche). To reduce proteins, DTT (final concentration 62.5 mM) was added to the 4x laemmli sample buffer (Bio-Rad). The membranes were blocked with 5% w/v milk in TBS/Tween 0.1% (TBS/T) and incubated with mouse monoclonal anti-StrepTagII (71590-3, EMD Millipore, 1:1000), rabbit polyclonal anti-His (236, CST, 1:1000), or rabbit polyclonal anti-SARS-CoV-2 Spike RBD (40592-T62, Sino Biological, 1:1000) antibodies in 2% BSA TBS/T (see **Table S1**). Secondary antibodies used were goat anti-mouse HRP (W4028, Promega, 1:2000) and donkey anti-rabbit HRP (Ab16284, Abcam, 1:2000) in 2% BSA TBS/T, respectively. Visualization of immunolabels was performed using West-Q Pico ECL solution (Gendepot), and chemiluminescent signals were captured using Amersham Hyperfilm (Cytiva, formerly GE Healthcare). The images were uniformly changed to gray scale.

### ELISA

MaxiSorp C-shaped 96 well plates were coated with Spike Ectodomain-Strep Trap, Spike Ectodomain-His Trap/SuperdexGF or Spike-RBD. Spike-RBD consists in 2.5 mg/mL (106.20 nM) of recombinant SARS-CoV-2 spike RBD protein from Sino Biologicals (40592-V08H). Spike Ectodomain-Strep Trap and Spike Ectodomain-His Trap/SuperdexGF are described above. For Spike Ectodomain-His Trap/SuperdexGF, 50 μg/mL (355.37 nM) was used, for Spike Ectodomain-Strep Trap, 2 μg/mL (14.21 nM) was used. The proteins were diluted in 50mM sodium carbonate (Na_2_CO_3_) - sodium bicarbonate (NaHCO_3_) coating buffer, pH 9.6, and allowed to bind to the well (50 microliters per well) for at 37°C 2 hours or at 4°C overnight. The coating mixture was removed from the wells and the wells were subsequently blocked with 5% non-fat dry milk in TBS (200 microliters per well) at room temperature for 1 hour. Serum samples were thawed on ice, serially diluted in TBS, and dispensed (50 microliters per well) in triplicates into the coated and blocked wells. For the performance assays shown in **Figure 3c-d**, 50 microliters of anti-StrepTagII antibody (**Table S1**, 1:500) in 2% BSA was added to the wells. The plate was incubated for at 37°C 1 hour, then washed three times with TBS, 0.1% Tween 20 (200 microliters per well). The wells were then incubated with horseradish peroxidase (HRP) conjugated secondary antibodies (**Table S1**, 50 microliters per well) at 37°C for 1 hour, and washed again three times with TBS, 0.1% Tween 20 (200 microliters per well). The last wash was removed from the well, and 3,3’,5,5’-Tetramethylbenzidine reagent (**Table S1**) was added to each well (50 microliters per well). The plate was incubated at room temperature for 15 minutes, and read at 650nm using the Versamax spectrophotometer from Molecular Devices. The reaction was stopped by adding 0.18 M sulfuric acid in each well (50 microliters per well), and the wells were read again at 450nm. In **Figures 4–6**, the negative control (NC) range represents the mean +/-standard deviation of all controls utilized. Controls included secondary antibody alone, and mixture of secondary antibodies and serially diluted serum samples on uncoated wells (high titers); these defined the range of negative controls (NC) signal. The OD450nm values were normalized (ratio) using the average OD450nm value of all 2017 healthy donor serum samples measured plus one standard deviation of this mean: OD450/[OD450 HD2017 + 1SD]. The highest titer was used for comparative analyses, which included the 1:50 serum dilution for the Spike Ectodomain-Strep Trap and the 1:100 serum dilution for the Spike-RBD. In some of the samples measured, the arbitrary cut off for positivity were normalized (ratio) OD450nm values greater than 1.5, borderline measurements were defined as normalized OD450nm of 1.2 to 1.5 or less, and OD450nm less than 1.2 were defined as negative.

**Figure 3:**
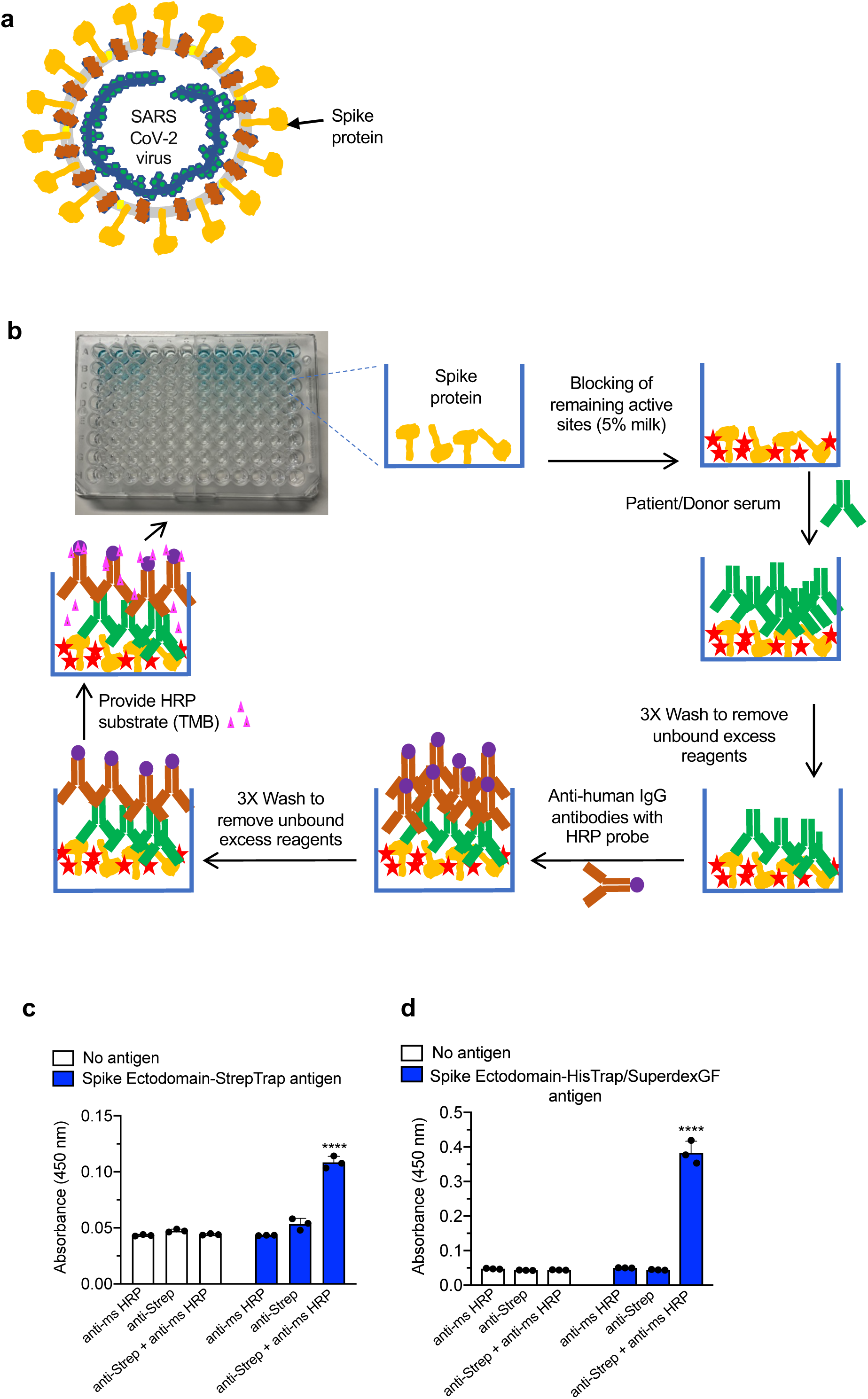
Design of ELISA-based test for spike protein. (**a**) Schematic of SARS-CoV-2 viral structure. Spike protein, orange; envelope protein, yellow; membrane glycoprotein, red; RNA, blue; nucleocapsid protein, green. (**b**) Schematic of ELISA protocol. Spike protein, orange; blocking reagent, red; primary antibody/serum/, green; secondary antibody, brown; horseradish peroxidase (HRP), purple; HRP substrate, magenta. (**c**) ELISA with Spike Ectodomain-StrepTrap antigen coating (2 μg/mL, 14.21 nM) and probed with anti-StrepTagII antibody. Absorbance at 450 nm reported. (**D**) ELISA with Spike Ectodomain HisTrap/SuperdexGF antigen coating (50 μg/mL, 355.37 nM) and probed with anti-StrepTagII antibody. Absorbance at 450 nm reported. Data is reported as mean ± standard deviation (s.d.) of 3 technical replicates for each sample. One-way ANOVA with Dunnett’s multiple comparison test, ^****^ *P* < 0.0001.

**Figure 4:**
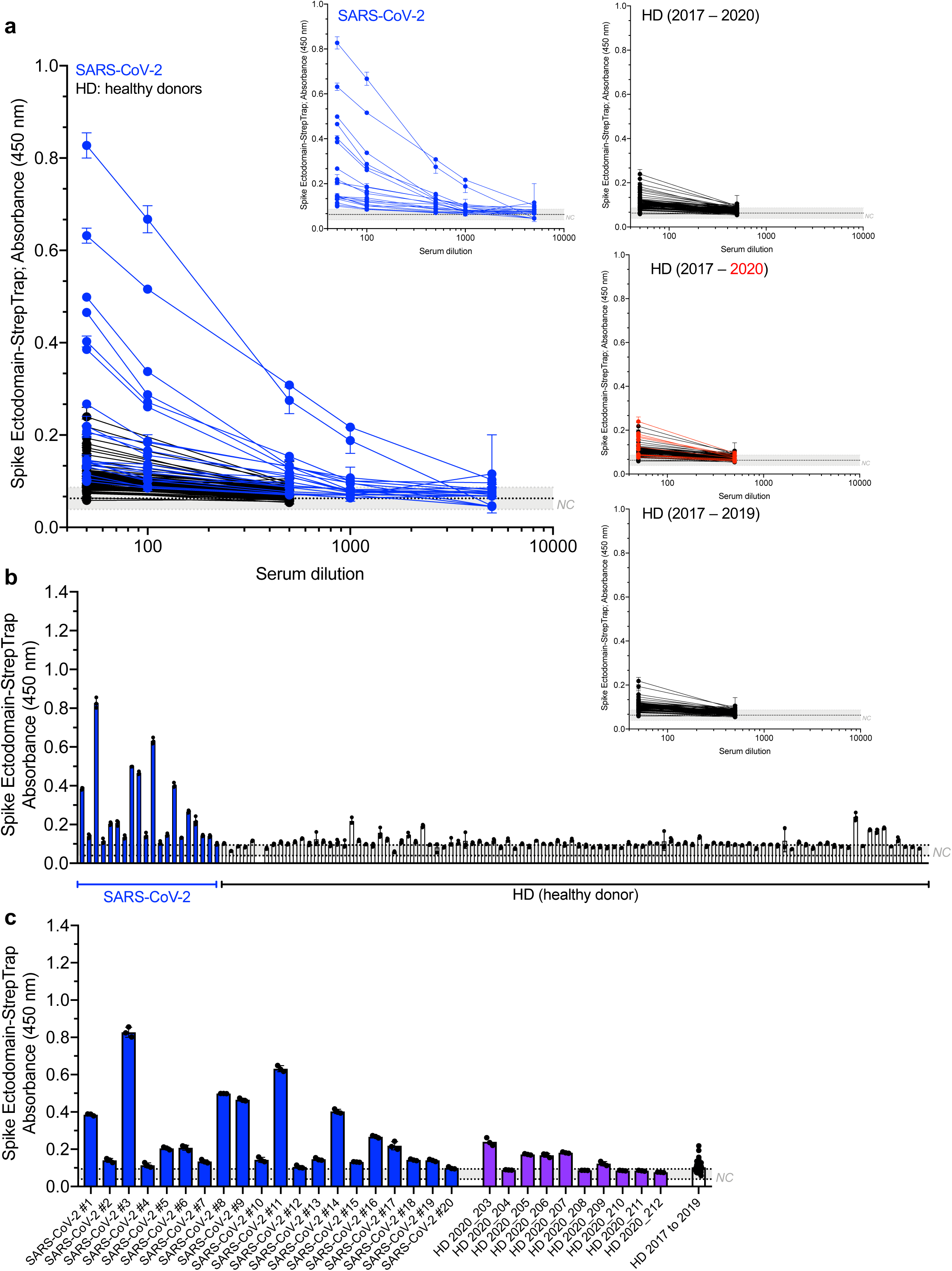
SARS-CoV-2 serological test with Spike Ectodomain-StrepTrap protein. (**a**) ELISA with Spike Ectodomain-StrepTrap protein coating and serial dilutions of serum. Fold dilution of serum indicated. Absorbance at 450 nm reported. SARS-CoV-2 (blue), N=20; healthy donors (HD, black), N=98. Inset graphs depict the data separated based on SARS-CoV-2 (left inset) or healthy donor serum collected from 2017 to 2020 (top right inset), 2017 to 2020 with 2020 samples depicted in red (middle right inset), and 2017 to 2019 (bottom right inset). Data is reported as mean ± standard deviation (s.d.) of 3 technical replicates for each sample. (**b**) ELISA with Spike Ectodomain-StrepTrap protein coating and 1:50 dilution of serum. Absorbance at 450 nm reported. SARS-CoV-2 (blue), N=20; healthy donors from 2017-2020 (HD, white), N=98. Data is reported as mean ± standard deviation (s.d.) of 3 technical replicates for each sample. (**c**) ELISA with Spike Ectodomain-StrepTrap antigen coating and 1:50 dilution of serum. Absorbance at 450 nm reported. SARS-CoV-2 (blue), N=20; healthy donors from 2020 (purple), N=10; healthy donors from 2017-2019 (white), N=88. Data is reported as mean ± standard deviation (s.d.) of 3 technical replicates for each sample for SARS-CoV-2 and HD 2020 samples. HD 2017–2019 data is presented with each dot representing the mean absorbance for a given serum sample diluted 1:50 and error bars depicting the s.d. of all samples. NC, negative control range, see methods.

**Figure 5:**
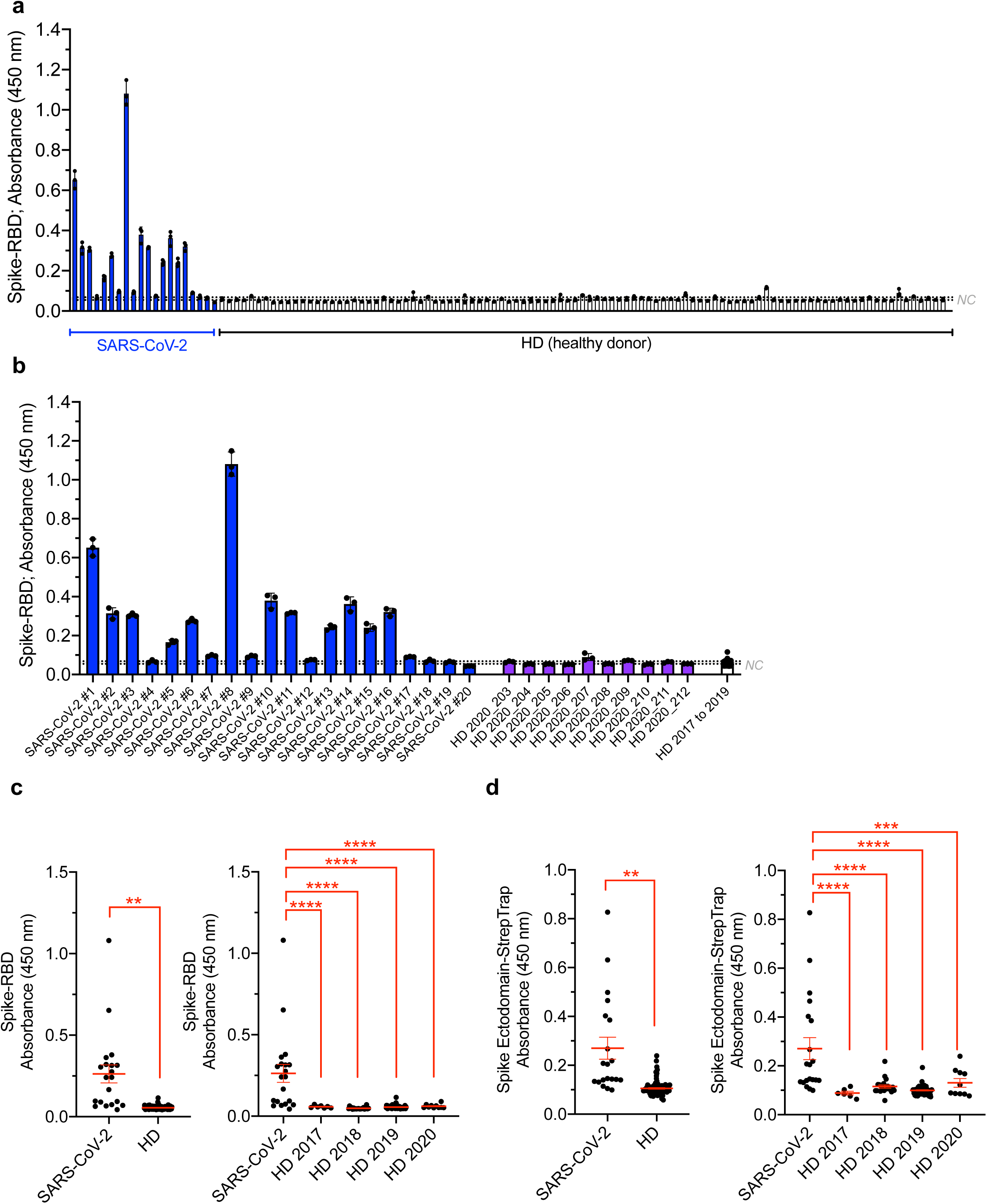
SARS-CoV-2 serological test with Spike-RBD protein. (**a**) ELISA with Spike-RBD protein coating and 1:100 dilution of serum. Absorbance at 450 nm reported. SARS-CoV-2 (blue), N=20; healthy donors from 2017-2020 (HD, white), N=99. Data is reported as mean ± standard deviation (s.d.) of 3 technical replicates for each sample. (**b**) ELISA with Spike-RBD antigen coating and 1:100 dilution of serum. Absorbance at 450 nm reported. SARS-CoV-2 (blue), N=20; healthy donors from 2020 (purple), N=10; healthy donors from 2017-2019 (white), N=89. Data is reported as mean ± standard deviation (s.d.) of 3 technical replicates for each sample for SARS-CoV-2 and HD 2020 samples. HD 2017-2019 data is presented with each dot representing the mean absorbance for a given serum sample diluted 1:100 and error bars depicting the s.d. of all samples. (**c**) Average 450 nm absorbance plotted for Spike-RBD antigen coating with SARS-CoV-2 and all healthy donors (left panel) or healthy donors separated based on time period of serum collection (right panel). Data is presented with each dot representing the mean absorbance for a given serum sample diluted 1:100. Welsh’s t-test (left panel) and one-way ANOVA with Dunnett’s multiple comparison test (right panel) performed with comparisons indicated. (**d**) Average 450 nm absorbance plotted for Spike Ectodomain-StrepTrap antigen coating antigen coating with SARS-CoV-2 and all healthy donors (left panel) or healthy donors separated based on time period of serum collection (right panel), related to Figure 4. Data is presented with each dot representing the mean absorbance for a given serum sample diluted 1:50, the red bar depicts the mean +/-standard error of the mean. Welsh’s t-test (left panel) and one-way ANOVA with Dunnett’s multiple comparison test (right panel) performed with comparisons indicated. ^**^ *P* < 0.01, ^***^ *P* < 0.001, ^****^ *P* < 0.0001.

**Figure 6:**
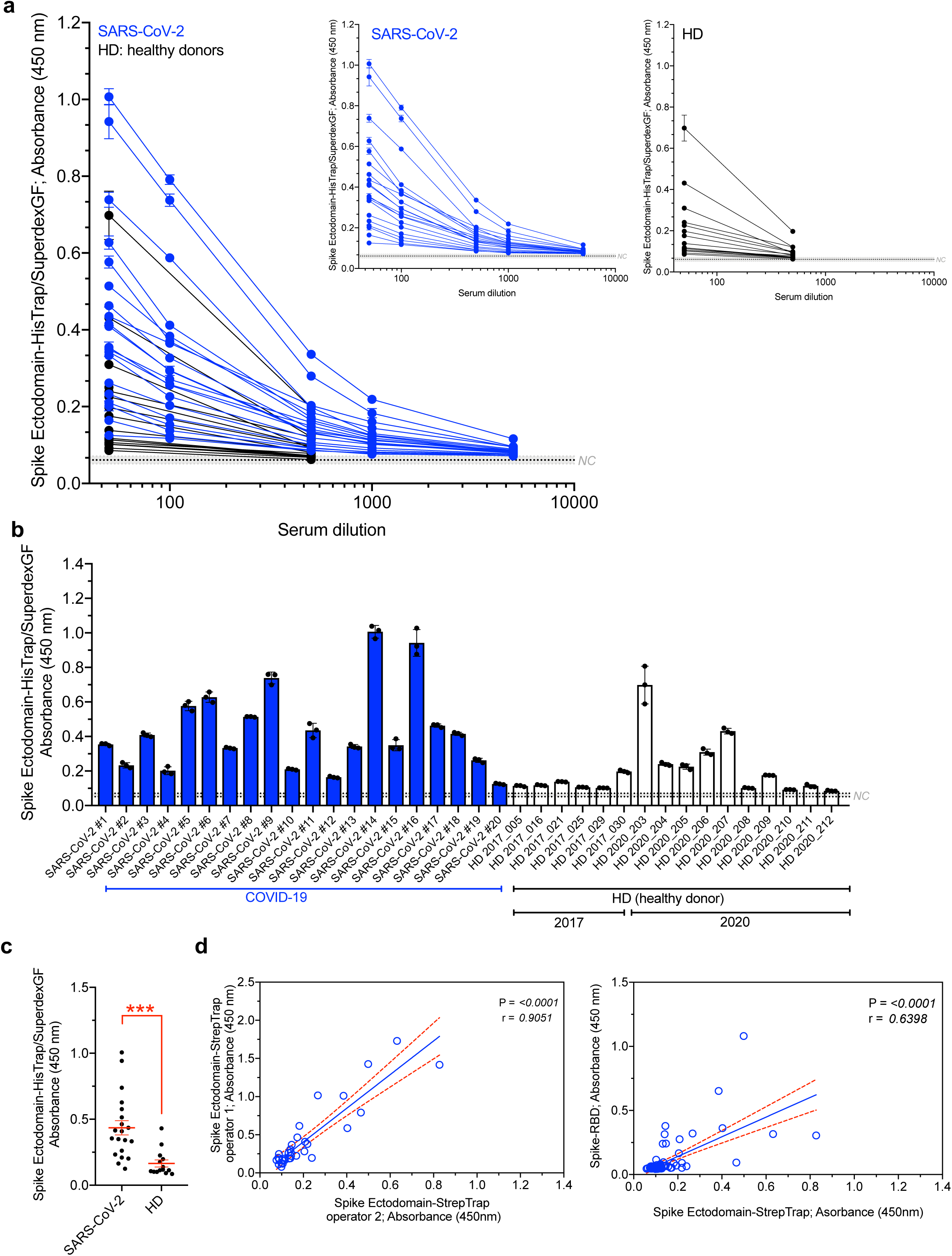
SARS-CoV-2 serological test with HisTrap/SuperdexGF purified spike protein. (**a**) ELISA with Spike Ectodomain-HisTrap/SuperdexGF antigen coating and serial dilutions of serum. Fold dilution of serum indicated. Absorbance at 450 nm reported. SARS-CoV-2 (blue), N=20; healthy donors (HD, black), N=16. Inset graphs depict the data separated based on SARS-CoV-2 (left inset) or healthy donors from 2017 and 2020 (right inset). NC, negative control range, see methods. Data is reported as mean ± standard deviation (s.d.) of 3 technical replicates for each sample. (**b**) ELISA with Spike Ectodomain-HisTrap/SuperdexGF antigen coating and 1:50 dilution of serum. Absorbance at 450 nm reported. SARS-CoV-2 (blue), N=20; healthy donors from 2017 and 2020 (HD, white), N=16. Data is reported as mean ± standard deviation (s.d.) of 3 technical replicates for each sample. (**c**) Average 450 nm absorbance plotted for Spike Ectodomain-HisTrap/SuperdexGF antigen coating with SARS-CoV-2 and all healthy donors. Data is presented with each dot representing the mean absorbance for a given serum sample diluted 1:50. SARS-CoV-2, N=20; healthy donors from 2017 and 2020 (HD), N=16. Welsh’s t-test performed. ^***^ *P* < 0.001. (**d**) Correlation between Spike-RBD and Spike Ectodomain-StrepTrap absorbance (left panel). Correlation between Spike Ectodomain-StrepTrap absorbance measured by two distinct operators (right panel). Pearson correlation coefficient (r) and *P* value reported.

### Data presentation and statistical analyses

Statistical analysis and generation of the graphs were carried out using GraphPad Prism. The mean and deviation from the mean presented in the figures are described in the figure legends.

## Results

### Production of SARS-CoV-2 recombinant proteins

The SARS-CoV-2 surface glycoprotein termed spike protein is composed of two subunits (S1 and S2); with the S1 subunit including the receptor binding domain (RBD) (**Figure 1a**). To generate SARS-CoV-2 spike proteins, the ectodomain of spike was expressed in 293T/17 cells using a previously described plasmid^21^ which was validated using restriction digestion (**Figure 1 b-c**). The expression product from this plasmid is termed ‘spike ectodomain’ (**Figure 1d** and^21^) with engineered Strep and His tags for subsequent purification from conditioned media of transfected cells. To generate the spike RBD, several plasmids that can yield modified RBD protein, including the His tagged secreted (Spike-RBD, **Figure 1e-f**) and the membrane-bound RBD using VSG-fusion protein were characterized (Spike-RBD-VSV-G, **Figure 1g-h**).

The spike proteins were purified from the conditioned media collected following transfection of 293T/17 cells (**Figure 2a**, see methods). The spike ectodomain was purified using a Strep trap (Spike Ectodomain-Strep Trap) or a His trap (Spike Ectodomain-His Trap/SuperdexGF, see methods). Quality of the purification of the Spike Ectodomain-Strep Trap was evaluated using FPLC chromatogram (**Figure 2b**), protein visualization following SDS-PAGE electrophoresis (**Figure 2c**), and western blot analysis for the immuno-detection of the His Tag and Strep Tag (**Figure 2d**). Spike RBD was validated using western blot and immuno-detection of the His-Tag and the RBD (**Figure 2e**). The chimeric Spike-RBD-VSV-G product was visualized by western blot analyses of transfected cell lysates against the Strep Tag (**Figure 2f**).

### Establishing ELISA for the detection of antibodies against SARS-CoV-2 proteins

The SARS-CoV-2 spike glycoprotein is found on the surface of the envelope of the virus (**Figure 3a**). Purified proteins are used to coat the ELISA plates, and the ELISA proceeds with sequential addition of serially diluted serum samples from patients (**Figure 3b**, see methods). The performance of the designed ELISA was tested using antibodies against the Strep tag found in both Spike Ectodomain-Strep Trap and Spike Ectodomain-His Trap/SuperdexGF. Anti-Strep antibodies specifically detected the Spike Ectodomain-Strep Trap (**Figure 3c**) and Spike Ectodomain-His Trap/SuperdexGF (**Figure 3d**) recombinant proteins.

### Detection of antibodies against SARS-CoV-2 proteins in human serum

Serial dilution of serum from healthy donors and patients with SARS-CoV-2 RT-PCR confirmed, hospitalized with COVID-19 infection were used to assess binding to the Spike Ectodomain-Strep Trap protein coated ELISA plates. Serum dilutions 1:50 and 1:100 showed elevated 450nm absorbance (A450nm) readings in 17 out of 20 of SARS-CoV-2 patients, whereas healthy donor serum showed A450nm readings within negative controls range (**Figure 4a-c**). We employed several negative controls to ensure reliability of our assays, including secondary only antibodies on coated wells, and mixture of secondary antibodies and serially diluted serum samples on uncoated wells. These defined the range of negative controls (NC) signal (see methods, **Figure 4**). Additional controls in assay development included uncoated wells with serum samples, coated wells alone, uncoated wells alone, and secondary antibodies on uncoated wells, all of which did not yield a signal above the background A450nm readings. Healthy donor serum (1:50 dilution) showed A450nm readings in the range of NC A450nm, whereas 4 out of 10 serum samples collected from January to February 2020 showed elevated A450nm readings (HD 2020, **Figure 4b-c**). Five out of 89 serum samples collected from 2017 to 2019 showed A450nm readings relatively higher than the NC A450nm readings, possibly accounting for cross reactivity of antibodies to other corona virus exposure (**Figure 4b**).

Serum samples evaluated against the Spike-RBD recombinant protein indicated detection of antibodies in 15 out of 20 SARS-CoV-2 positive patients (**Figure 5a**). A450nm readings were not elevated in all but one serum sample of the healthy donors collected in 2020, and in one of the healthy donors collected in 2019 (**Figure 5b**). There was no detection of Spike-RBD antibodies in 5 out of 20 SARS-CoV-2 patient serum, and 3 out of 20 were borderline (**Figure 5b**). The A450nm readings were significantly elevated in the serum of the SARS-CoV-2 patients compared to all healthy donor serum (**Figure 5c-d**).

To further confirm the SARS-CoV-2 spike protein antibody detection using the purified Spike Ectodomain-Strep Tag protein, a subset of serum samples was also tested against the Spike Ectodomain-His Trap/SuperdexGF protein (**Figure 6**). Similar to the results obtained with Spike Ectodomain-Strep Tag, elevated A450nm were noted in SARS-CoV-2 serum samples, as well as in some of the 2020 healthy donor serum (e.g. # 203, **Figure 6a-b**). The A450nm readings were significantly elevated in the serum of the SARS-CoV-2 patients compared to healthy donor serum (**Figure 6c**).

Additional internal validation of the ELISA designed in this study included a two-independent-operator analysis of serum samples against the Spike Ectodomain-Strep Tag protein. Each operator followed a defined protocol without prior consultation regarding assay logistics. The A450nm readings indicated robust correlation between the two operators (**Figure 6d**), supporting a reproducible methodology and protocol definition. The A450nm readings obtained using the Spike-RBD protein (**Figure 5**) and the Spike Ectodomain-Strep Tag (**Figure 3**) also significantly correlated with one another, with most healthy donor samples exhibiting lack of antibody detection (**Figure 6d**).

### Antibodies in SARS-Cov-2 patient serum correlation with clinical progression

To compare relative positivity for antibodies against the ectodomain of Spike and RBD, the normalized ratio of A450nm was used (**Figure 7a**). Specifically, the averaged A450nm reading from the 2017 healthy donors plus one standard deviation of this mean was used as the denominator (see methods). The heat map of the computed ratios showed elevated ratios for the serum of the SARS-CoV-2 patients and healthy donors serum collected in 2020. Arbitrary cut-off points were used to aid the visual distribution of the positive and negative serum. Ratios greater than 1.5 were labeled positive, ratios between 1.2 and 1.5 were labeled borderline, and ratios less than 1.2 were labeled negative (**Figure 7b**). With these cut-off points, 10 SARS-CoV-2 serum samples were positive, 7 were borderline, and 3 were negative (n=20 total) for antibody detection against the Spike Ectodomain-Strep Tag protein (**Figure 7b**). Twelve SARS-CoV-2 serum samples were positive, 3 were borderline, and 5 were negative (n=20 total) for antibody detection against the Spike-RBD protein (**Figure 7b**). Three SARS-CoV-2 serum samples consistently negative for antibodies against both Spike Ectodomain-Strep Tag and Spike-RBD protein, and 2 of the SARS-CoV-2 serum samples negative for antibodies against Spike-RBD protein while borderline for antibodies against Spike Ectodomain-Strep Tag protein (**Figure 7b**). In healthy serum samples collected in 2020, 4 out of 10 serum samples were positive for antibodies against both Spike Ectodomain-Strep Tag, amongst which 1 sample was also positive for antibodies against Spike-RBD protein (**Figure 7b**).

**Figure 7:**
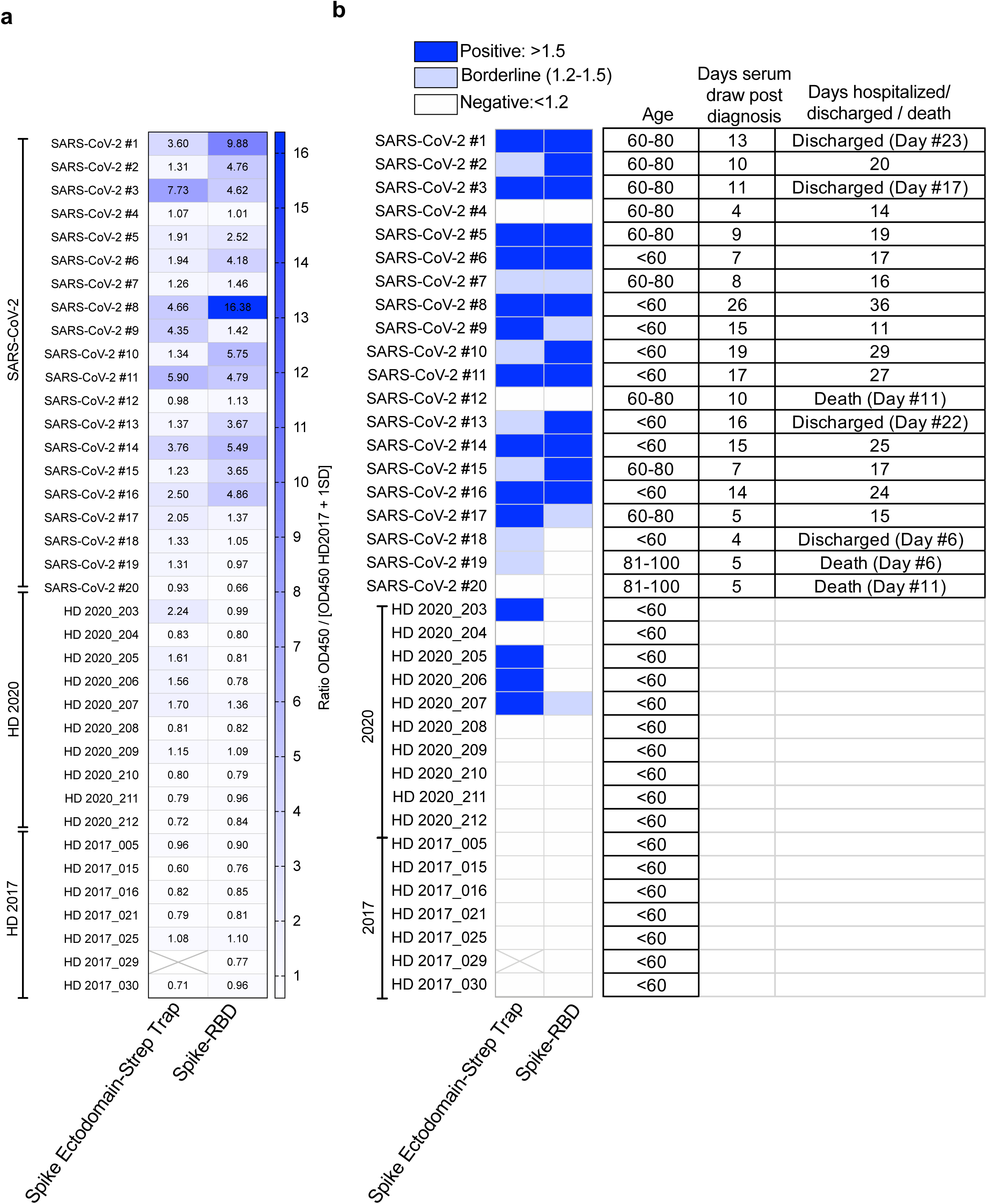
Association between detection of serological spike protein IgG and clinical features. (**a**) Heatmap of relative OD450 levels of SARS-CoV-2, HD from 2020, and HD from 2017. Data is represented as the ratio of OD450 to [mean OD450 of HD 2017 + one standard deviation (s.d.)]. Ratio values are indicated in heatmap, see also **Table 1**. (**b**) Positive, borderline, and negative cut-off label for IgG to Spike Ectodomain-StrepTrap or RBD, see also **Table 1**.

SARS-CoV-2 serum samples positive for antibodies against Spike-RBD protein (n=12) did not appear to correlate with recovery (shorter hospitalization or recovery). Three out of the 12 COVID-19 positive cases were discharged while the other 9 remain hospitalized (**Figure 7b, Table 1**). Three of the 4 SARS-CoV-2 patients with no detectable (negative) antibodies against Spike-RBD protein died, and the one recovered and was discharged (**Figure 7b, Table 1**). Notably, two of the 3 negative SARS-CoV-2 patients that died were between 81 and 100 years of age, corroborating age as a critical morbidity factor in SARS-CoV-2 infection outcome (**Figure 7b, Table 1**). Our results do not support that detection of serum antibodies against SARS-CoV-2 spike proteins was associated with rapid recovery or lack of comorbidities (**Figure 7b, Table 1**), although a larger cohort study is required.

## Discussion

We report the detection of circulating IgG antibodies against SARS-CoV-2 spike protein and RBD in patients with symptomatic, SARS-CoV-2 virus confirmed, hospitalized COVID-19 patients. Some of the patients lacked antibodies against spike RBD despite showing detection of antibodies to the spike ectodomain. This suggests that some patients can launch polyclonal IgG response to the 170k Da glycoprotein but not to the RBD that is responsible for the binding to the ACE2 receptors epithelial cells. Our study also indicates that while three patients that recovered and were discharged from the hospital had antibodies to RBD, one of them did not. Despite a very small sample size, our results highlight that individuals with non-RBD antibodies can recover from SARS-CoV-2 infection. Whether antibodies to other regions of spike glycoproteins can inform on neutralization or immunity remains to be tested.

The detection of antibodies against both the spike glycoprotein and the RBD was not associated with a measurable decrease in recovery time, compared to patients who did not exhibit antibody response. Some patients exposed to SARS-CoV-2 likely developed antibodies to spike glycoprotein and RBD but still progressed to severe COVID-19 requiring intensive care.

The development of ELISA assays to detect circulating IgG antibodies against SARS-CoV-2 proteins will likely serve the research community to advance our understanding of the COVID-19 disease. It may be valuable to distinguish whether antibodies to distinct regions of spike antigen, low titers, or complete lack of antibody detection, is associated with co-morbidities recently associated with COVID-19, namely hypertension, obesity, and diabetes^9^. The complete lack of antibody detection despite apparent recovery may suggest distinct immune mechanisms to overcome the virus, as reported for the role T cells and the lack of antibodies from B cells in SARS-CoV immunity^6^. Such new knowledge may inform on new therapeutic avenues.

Detection of circulating SARS-CoV-2 antibodies in healthy donors whose serum was collected earlier this year (February 2020), suggest the possibility that our communities may have already been exposed to the virus at much higher rates than anticipated, with asymptomatic outcome. Community transmission of SARS-CoV-2 in Wuhan (China) is now estimated to have begun in early January 2020^22^, supporting potential cases of COVID-19 in the US population as early as January 2020. Testing of healthy donors, with and without prior exposure will be essential to address this knowledge gap, and this need may be uniquely addressed by research communities.

## Data Availability

The data listed with the manuscript, any additional information can be requested to the corresponding author.

## Acknowledgements

This work was primarily supported by funds from MD Anderson Cancer Center to Dr. Kalluri. We thank Lisa Becker for her help in performing ELISAs, and Jackie Parchem and Michelle Kirtley for administrative support. We thank Dr. McLellan of University of Texas Austin for the spike ectodomain construct.

---

**Table S1.**
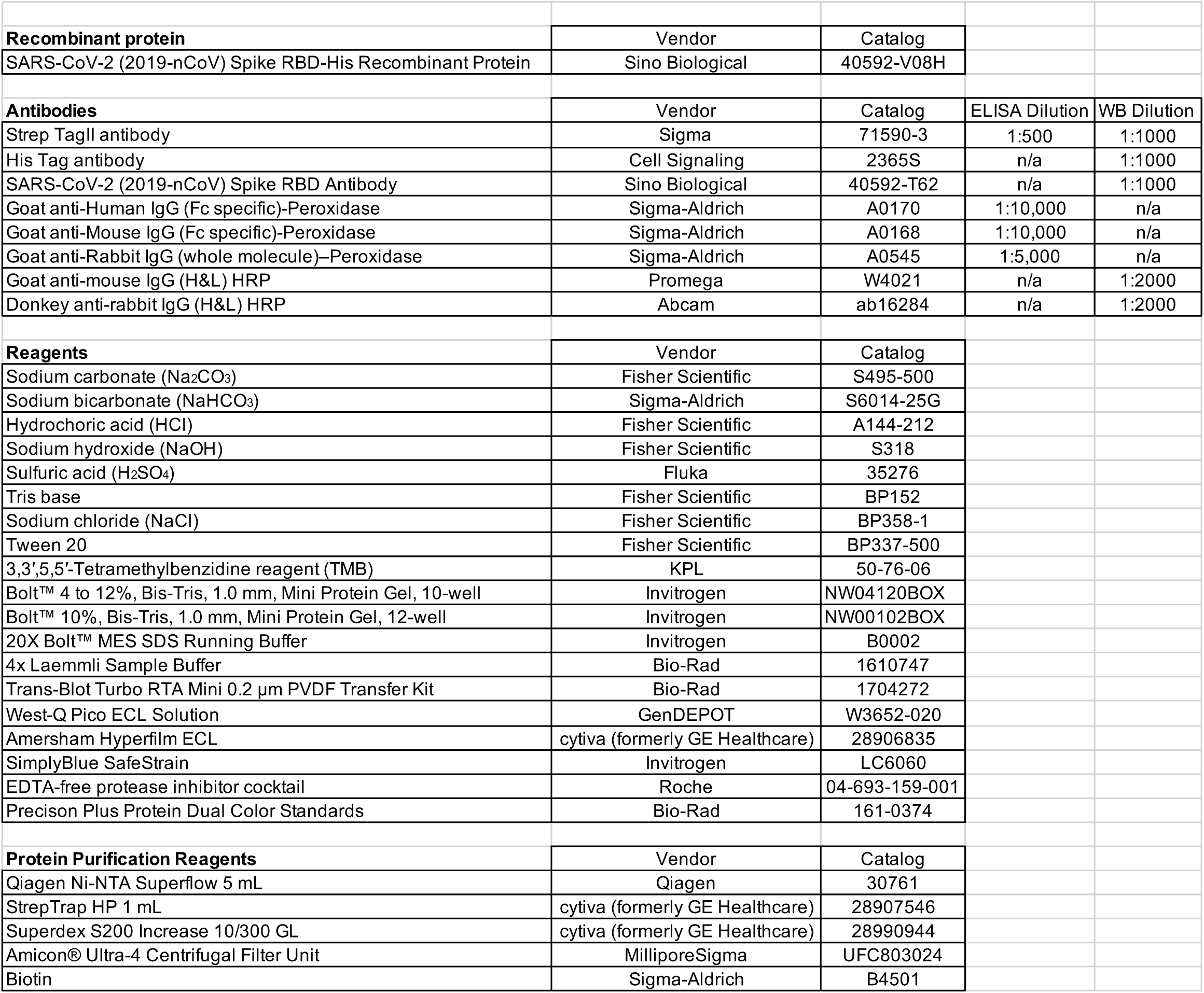
Reagents.

